# Developing and Validating a Computable Phenotype for the Identification of Transgender and Gender Nonconforming Individuals and Subgroups

**DOI:** 10.1101/2020.08.04.20168161

**Authors:** Yi Guo, Xing He, Tianchen Lyu, Hansi Zhang, Yonghui Wu, Xi Yang, Zhaoyi Chen, Merry Jennifer Markham, François Modave, Mengjun Xie, William Hogan, Christopher A. Harle, Elizabeth A Shenkman, Jiang Bian

## Abstract

Transgender and gender nonconforming (TGNC) individuals face significant marginalization, stigma, and discrimination. Under-reporting of TGNC individuals is common since they are often unwilling to self-identify. Meanwhile, the rapid adoption of electronic health record (EHR) systems has made large-scale, longitudinal real-world clinical data available to research and provided a unique opportunity to identify TGNC individuals using their EHRs, contributing to a promising routine health surveillance approach. Built upon existing work, we developed and validated a computable phenotype (CP) algorithm for identifying TGNC individuals and their natal sex (i.e., male-to-female or female-to-male) using both structured EHR data and unstructured clinical notes. Our CP algorithm achieved a 0.955 F1-score on the training data and a perfect F1-score on the independent testing data. Consistent with the literature, we observed an increasing percentage of TGNC individuals and a disproportionate burden of adverse health outcomes, especially sexually transmitted infections and mental health distress, in this population.

## Introduction

As a health disparity population designated by the National Institutes of Health (NIH), sexual and gender minority (SGM) individuals, especially transgender and gender nonconforming (TGNC) people, face a disproportionate burden of adverse health outcomes.^1^ Although there is a growing body of literature on the unique health issues among the TGNC population, they remain severely underserved and existing data on TGNC health are scarce. Under-reporting of TGNC status is common since TGNC individuals are often unwilling to self-identify and participate in traditional surveys due to issues related to social and economic marginalization, stigma, and discrimination, leading to challenges in obtaining population-based estimates. Because the SGM population represents a relatively small proportion of the population, it is labor-intensive and costly to recruit a large enough sample in general population surveys for meaningful analysis of the SGM population and their subgroups.^2^

The last few years have witnessed a rapid adoption of electronic health record (EHR) systems in the United States (US) and these systems have become an integral part of the health care system. As of 2017, 85.9% (nearly 9 in 10) of office-based physicians had adopted an EHR system, and 79.7% (nearly 4 in 5) had adopted a certified EHR system that meets the requirements of the US Department of Health and Human Services.^3^ Likewise, 96% of hospitals utilized an EHR in 2017.^4^ Furthermore, the widespread adoption of EHR systems has led to the creation of large-scale national and international clinical data research networks. The national Patient-Centered Clinical Research Network (PCORnet) funded by the Patient-Centered Outcomes Research Institute (PCORI) is one of the most prominent examples. PCORnet is a “*network of networks*” currently containing data for over 66 million patients collected through 348 health systems in the US.^5^ For example, the OneFlorida Clinical Research Consortium is one of the 9 clinical data research networks (CDRNs) contributing to the PCORnet.^6^ OneFlorida includes 12 unique healthcare organizations providing care for over half of all Floridians (~15 million) through 914 clinical practices and 22 hospitals that cover all 67 counties in Florida.

The widespread adoption of EHRs and the creation of clinical research networks with large collections of EHRs have made large-scale, longitudinal clinical data available for research. The FDA recently coined the terms real-world data (RWD) to refer to information derived from sources outside research settings, including EHRs, claims, and billing data among others. Still, a key to successfully using these RWD in health disparities and outcomes research is the ability to accurately identify the populations of interest. EHRs contain not only important structured data, such as diagnoses and procedures, but also unstructured clinical narratives such as physician’s notes. More than 80% of the clinical information in EHRs is documented in clinical narratives,^7^ which often contain more detailed patient information including TGNC status. In a previous study, Roblin *et al* used a combination of keywords (e.g., “*transgender*”) and relevant diagnostic codes and identified 271 possible TGNC individuals out of 813,737 members in the Kaiser Permanente Georgia EHR system.^8^ Of the 271 possible TGNC individuals, 185 (68%) were confirmed through manual chart review.^8^ In a follow up study, Quinn *et al* followed the same approach and defined a Study of Transition Outcomes and Gender (STRONG) cohort to assess health status of TGNC individuals.^9^ They used data from Georgia, Northern California and Southern California Kaiser Permanente health plans and identified 12,457 potential TGNC individuals. Of these, 6,456 (52%) were confirmed through manual chart review.^9^ There are a few key gaps in these two existing studies: (1) they only considered International Classification of Diseases, Ninth Revision, Clinical Modification (ICD-9-CM) codes, as only EHR data from before 2014 were available in their studies; (2) they used internal Kaiser Permanente codes in combination with ICD-9 V codes that are not generalizable to other health systems; (3) the keywords for identifying potential TGNC individuals from clinical notes were limited and missing important potential keywords (e.g., “*trans male*”, “*trans female*”); (4) they did not consider the discrete gender identity (i.e., the 2011 IOM report made recommendations to gather sexual orientation and gender identity [SO/GI] data in EHRs as part of the meaningful use objectives;^2^ and the Centers for Medicare & Medicaid Services [CMS] set the final rule in 2015 to require sexual orientation and gender identity fields for meaningful use stage 3 certification^10^); and (5) they used two trained reviewers to manually review deidentified text strings for all unconfirmed cases, where a more automated approach is needed to scale to data from other health systems.

Using diagnostic codes alone (i.e., ICD-9/10-CM) has poor specificity and sensitivity for cohort identification in EHRs. This is why computable phenotypes (i.e., “*clinical conditions, characteristics, or sets of clinical features that can be determined solely from EHRs and ancillary data sources and does not require chart review or interpretation by a clinician*.”^11^) are needed.^12,13^ In this study, we aimed to build upon Quinn et al’s work^9^ to develop and validate a computable phenotype (CP) for accurate and automated identification of TGNC individuals, their subgroups (i.e., male-to-female, female-to-male), and their natal sex (i.e., male or female) using both structured and unstructured data from an academic health center (i.e., the University of Florida Health [UF Health]). We assessed the prevalence of TGNC in UF Health and describe the general health status (e.g., chronic diseases) of the identified TGNC cohort. Resources from this study, such as the list of diagnostic and procedure codes as well as the keywords, are available at https://github.com/bianjiang/tgnc_ehr_computable_phenotype.

## Methods

### Data source

With a protocol approved by the UF Institutional Review Board (IRB), we obtained individual-level patient data from the UF Health Integrated Data Repository (IDR), a clinical data warehouse (CDW) that aggregates data from UF’s various clinical and administrative information systems, including the Epic EHR system. The UF IDR contains demographics, clinical encounter data, diagnoses, procedures, lab results, medications, select nursing assessments, co-morbidity measures and select perioperative anesthesia information system data. As of January 2020, the IDR contains records of 1.2 million patients with over 1 billion observation facts.^14^

### Overall study design

***Figure 1*** shows the cohort ascertainment diagram of our approach to identifying TGNC individuals and their natal sex based on both structured and unstructured EHR data. We used a 3-step process to develop the CP for TGNC: (1) ***Step 1***—search EHRs to identify potential TGNC individuals based on the discrete gender identity field, relevant diagnosis and procedure codes, and relevant TGNC keywords; (2) ***Step 2***—validate the cohort through manual chart review of selected samples to derive CP rules; and (3) ***Step 3***—identify CP rules for determining TGNC subgroups and natal sex assignment (i.e., transfeminine/male-to-female [MTF] vs. transmasculine/female-to-male [FTM]).

**Figure 1.**
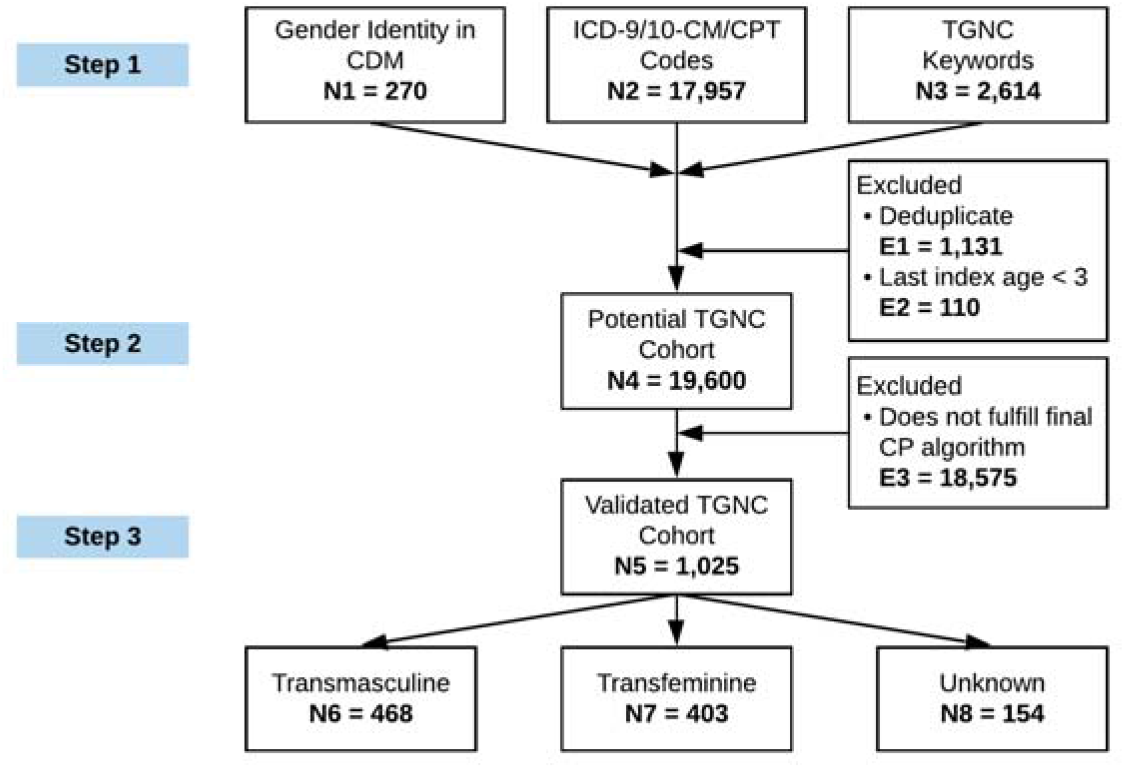
Cohort ascertainment diagram for identifying transgender and gender nonconforming (TGNC) individuals and their natal sex.

#### Step 1: Search EHRs to identify potential TGNC individuals

We used an iterative process to retrieve data from the UF IDR considering three different scenarios for identifying potential TGNC individuals. First, it was straightforward to include patients whose (1) structured gender identity fields were recorded as “t*ransgender female / male-to-female*”, “*transgender male / female-to-male*”, “*nonbinary*”, and “*other*”; and (2) gender identity fields had different values from the same patients’ sex fields. Second, we adopted the diagnostic and procedure codes used in Quinn et al but with significant expansion based on online resources such as the “*Gender Reassignment Surgery Model National Coverage Determination (NCD)*” from the Transgender Medicine Model NCD Working Group and the NCD for gender dysphoria and gender reassignment surgery from the Centers for Medicare & Medicaid Services (CMS),^15^ where common Current Procedural Terminology (CPT) and ICD-9/10-CM codes were listed for gender reassignment surgery. Third, we expanded and refined the keywords that could be used to identify potential TGNC individuals iteratively, using the clinical narratives in their EHRs based on (1) Quinn et al’s work^9^, (2) our prior work^16^ on identifying gender identification terms using social media data, (3) other online resources such as those from the Fenway Health’s glossary of gender and transgender terms^17^, and (4) sample notes from the UF IDR that contained any of these keywords, until no new keywords were found. **Table 1** summarizes the three search strategies to identify potential TGNC individuals. We retrieved both structured EHR data (e.g., encounter data, diagnoses, procedures, etc.) and unstructured clinical narratives (e.g., progress note, discharge summary, radiology reports, etc.) for all patients in the “*Potential TGNC*” cohort.

**Table 1.** Initial search criteria used to identify potential TGNC individuals from EHR data.

|  | Value Sets/Codes/Keywords |
| --- | --- |
| **Gender Identity (GI)** |  |
| GI as TGNC | “transgender female / male-to-female”, “transgender male / female-to-male”, “nonbinary”, and “other” |
| GI/SEX Conflict | GI recorded as ‘male’ with SEX as ‘female’ or GI recorded as ‘female’ with SEX as ‘male’ |
| **Diagnosis Codes (Selected Examples)** |  |
| ICD-9-CM | e.g., 302.3 – “Transvestic fetishism”, 302.5 – “Trans-sexualism”, 302.85 – “Gender identity disorder in adolescents or adults”, etc. |
| ICD-10-CM | e.g., F64 - “Gender identity disorders”, F65.1 – “Transvestic fetishism”, Z87.890 – “Personal history of sex reassignment”, Z90.72 – “Acquired absence of ovaries”, Z90.79 – “Acquired absence of other genital organ” |
| **Procedure Codes (Selected Examples)** |  |
| CPT | e.g., 58720 – “Salpingo-oophorectomy”, 58571 – “Laparoscopy, surgical with total hysterectomy”, etc. |
| **Keywords (Selected Examples)<sup>a</sup>** |  |
|  | e.g., “transgender”, “transsexual”, “male to female”, “female to male”, “trans female”, “trans male”, “nonbinary”, “gender nonconforming”, “gender nonconformity”, “preferred pronoun”, “gender dysphoria”, “gender disorder”, “sex reassignment”, “gender reassignment”, etc. |
| <sup>a</sup>Note that keyword searches were NOT case sensitive; and we also considered variations of the keywords (e.g., “trans male”, “transmale”, “trans-male”). |  |

#### Step 2: Validate the cohort through manual chart review of selected samples to derive CP rules

Based on all the prior work and discussions with clinicians who provide care to TGNC individuals, we started with 6 initial rules: having a GI value indicative of TGNC status, having conflicting values between the GI and Sex fields, having 1 of the relevant diagnostic codes, having more than 1 relevant diagnostic codes, having 1 or more of the procedure codes, and having at least 1 of the relevant keywords. We excluded patients whose age was less than 3 based on Quinn’s work. We excluded patients only having hits with zero precision keywords (i.e., keywords that did not yield any positive cases in our samples): “*gender identity*”, and “*identified as*”, as these keywords often exist in templated clinical note as field labels. As shown in **Table 2**, for our training set, we randomly selected samples from 25 different combinations of these 6 rules for manual chart review.

**Table 2.**
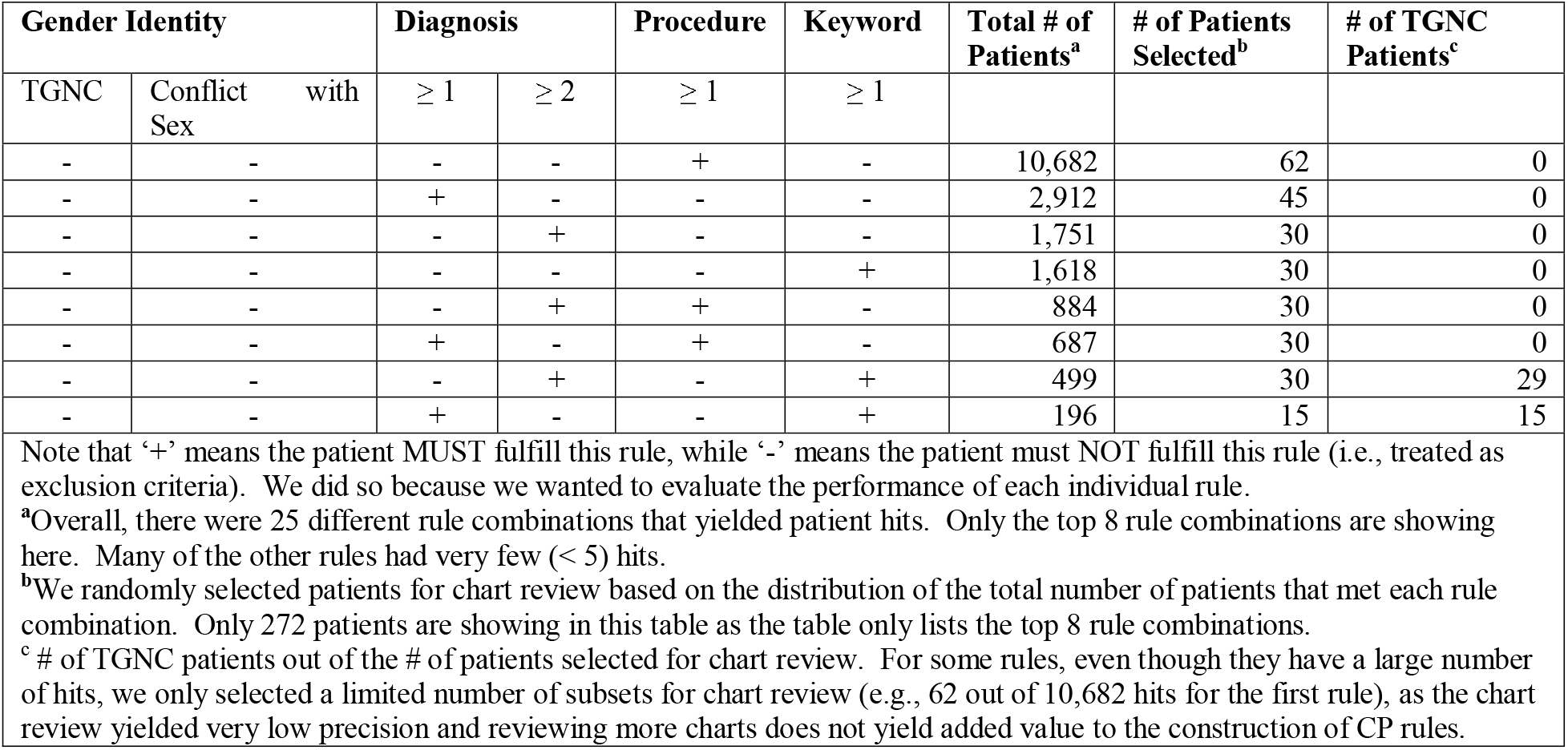
Different combinations of the 6 base rules, the number of patients met the criteria for each rule combination, the number of randomly selected samples for manual chart reviews.

For the rule contingent on relevant keywords, we also evaluated the precision as false positives (i.e., containing a TGNC related keyword but was not truly indicative of TGNC status) exist for three main reasons: (1) the keyword was ambiguous (e.g., “*transvestite life style*”), (2) the sentence containing the keyword was a negation (e.g., “*In regard to the gender issue, it can be appropriate for girls and boys to play with cross gender toys. Diagnosis with a Gender Identity Disorder is not valid nor helpful in this ca*se.”), and (3) the recorded keyword was referring to individuals other than the patient (e.g., “*Her sibling has transitioned from female to male*.”).

#### Step 3: Identify CP rules for determining TGNC subgroups and natal sex assignment

Each identified TGNC individual was further categorized as (1) transfeminine (i.e., male-to-female [MTF]), (2) transmasculine (i.e., female-to-male [FTM]), or (3) unclear, primarily based on the existence of relevant keywords (e.g., “*male-to-female*”) or their variations (e.g., “*male to female*”, “*m2f*”). During the manual chart review, the reviewers were instructed to identify each TGNC individual’s natal sex: ‘*male’*, ‘*female’*, or ‘*unclear’*. Built upon Quinn’s work, we determined the TGNC individual’s natal sex assignment based on (1) clinical notes that contain natal sex anatomy (e.g., ‘*testes’*, ‘*ovaries’*), (2) history of specific procedures (e.g., orchiectomy or hysterectomy identified in both structured data using CPT codes and in clinical notes through searching for these procedures), and (3) evidence of hormonal therapy (e.g., estrogen or testosterone identified in both structured medication lists and in clinical notes). **Table 3** shows the procedures codes (i.e., CPT), medication codes (i.e., RxNorm), and keywords used to determine MTF or FTM status of an TGNC individual.

**Table 3.**
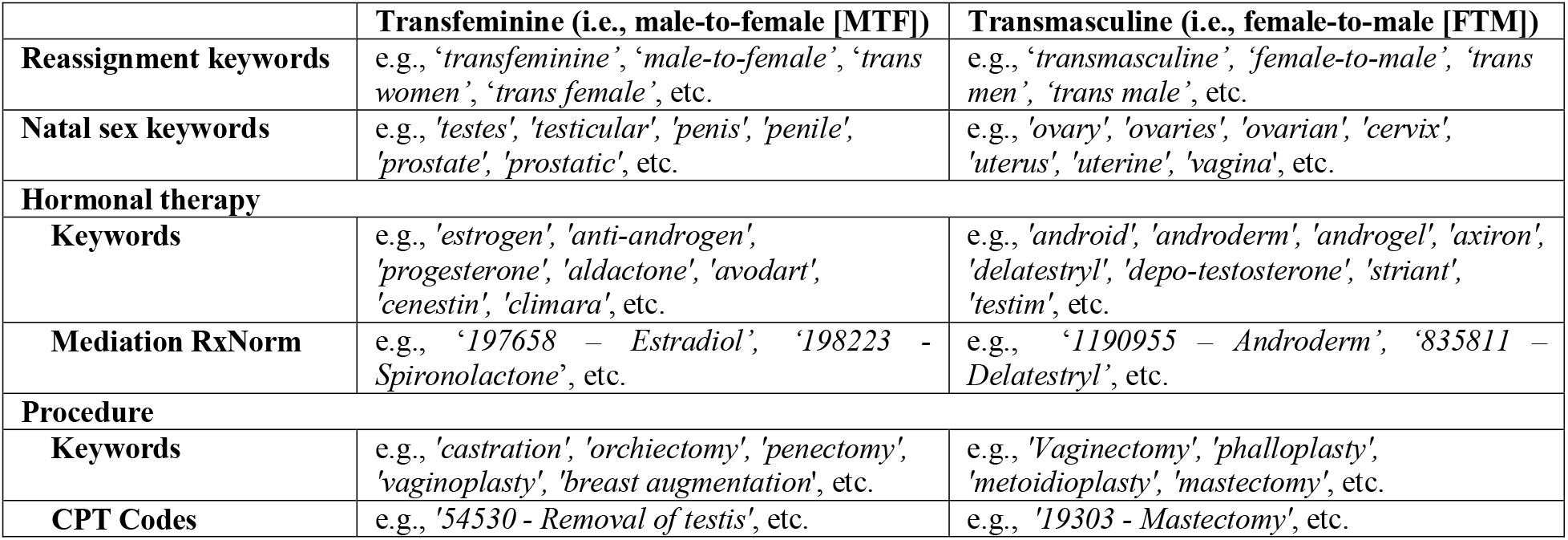
Example procedure codes, medication codes, and keywords used to determine natal sex assignment.

|  | Transfeminine (i.e., male-to-female [MTF]) | Transmasculine (i.e., female-to-male [FTM]) |
| --- | --- | --- |
| **Reassignment keywords** | e.g., ‘*transfeminine*’, ‘*male-to-female*’, ‘*trans women*’, ‘*trans female*’, etc. | e.g., ‘*transmasculine*’, ‘*female-to-male*’, ‘*trans men*’, ‘*trans male*’, etc. |
| **Natal sex keywords** | e.g., ‘*testes*’, ‘*testicular*’, ‘*penis*’, ‘*penile*’, ‘*prostate*’, ‘*prostatic*’, etc. | e.g., ‘*ovary*’, ‘*ovaries*’, ‘*ovarian*’, ‘*cervix*’, ‘*uterus*’, ‘*uterine*’, ‘*vagina*’, etc. |
| **Hormonal therapy** |  |  |
| **Keywords** | e.g., ‘*estrogen*’, ‘*anti-androgen*’, ‘*progesterone*’, ‘*aldactone*’, ‘*avodart*’, ‘*cenestin*’, ‘*climara*’, etc. | e.g., ‘*android*’, ‘*androderm*’, ‘*androgel*’, ‘*axiron*’, ‘*delatestryl*’, ‘*depo-testosterone*’, ‘*striant*’, ‘*testim*’, etc. |
| **Mediation RxNorm** | e.g., ‘197658 – Estradiol’, ‘198223 – Spironolactone’, etc. | e.g., ‘1190955 – Androderm’, ‘835811 – Delatestryl’, etc. |
| **Procedure** |  |  |
| **Keywords** | e.g., ‘*castration*’, ‘*orchiectomy*’, ‘*penectomy*’, ‘*vaginoplasty*’, ‘*breast augmentation*’, etc. | e.g., ‘*Vaginectomy*’, ‘*phalloplasty*’, ‘*metoidioplasty*’, ‘*mastectomy*’, etc. |
| **CPT Codes** | e.g., ‘54530 - Removal of testis’, etc. | e.g., ‘19303 - Mastectomy’, etc. |

### Evaluation and descriptive of cohort characteristics

Following best practice in developing computable phenotypes, we randomly selected 300 sample charts based on the distribution of the available data by the initial rule combinations as shown in ***Table 2***. An initial annotation guideline was first developed by the study team, and two annotators (XH and TL) independently reviewed the first 20 samples. The inner-rater agreement between the two annotators achieved a Cohen’s kappa of 0.88 during the initial round of review. Conflicts between the two annotators were resolved through discussions with the entire study team. The annotation guideline was revised and updated iteratively. The two annotators subsequently annotated the remaining training samples independently, and they were instructed to be conservative when a case was deemed uncertain. Cases with conflicting results were discussed and resolved with a third reviewer (JB). Based on these 300 training samples, a set of CP rules were derived. Lastly, the two annotators annotated another 100 randomly selected samples, which served as an independent testing set. We evaluated and reported the performance of (1) individual CP rules, (2) a CP algorithm considering CP rules with only structured data, and (3) a CP algorithm considering both structured and unstructured EHR data, in terms of specificity, sensitivity, positive predictive value (PPV), negative predictive value (NPV), and F1-score on both the training and testing sets.

## Results

### Development of the computable phenotype for the identification of TGNC

As shown in ***Figure 1***, we identified 19,600 potential TGNC individuals from the UF Health clinical data warehouse (i.e., the UF Health IDR) after deduplicating the records (i.e., 1,131 duplicates) across the three initial criteria indicative of TGNC status (i.e., the discrete GI field, relevant diagnosis and procedure codes, and relevant TGNC keywords, as shown in ***Table 1***) and excluding patients whose were younger than three (i.e., 110 patients).

To create a training dataset, we selected 300 patients from the potential TGNC individuals according to the distribution of patients in each of the rule combinations as shown in ***Table 1***. Within each rule combination, the selection was random. We then selected another simple random sample of 100 potential TGNC individuals as a hold-out testing dataset. Out of the 300 training samples, 68 were confirmed TGNC individuals and 44 individuals’ TGNC status could not be confirmed due to lack of sufficient data. Out of the 68 TGNC individuals, 27 were transfeminine/MTF and 41 were transmasculine/FTM. In the 100 testing samples, 6 individuals had insufficient data for determining their TGNC status, while 5 individuals were confirmed as TGNC (i.e., 1 transfeminine/MTF and 4 transmasculine/FTM).

Using the manual chart review results as the gold-standard, we first evaluated the performance of individual CP rules (as shown in ***Table 1***) considering two scenarios: (1) structured data only, and (1) both structured and unstructured data. ***Table 4*** shows the individual CP rules with the best performance in terms of specificity, sensitivity, PPV, NPV, and F1-score under the two scenarios. To objectively measure the performance of individual CP rules, we made sure a patient was classified as a TGNC if and only if the patient met the inclusion criteria (‘+’ in ***Table 1***) and did not meet the exclusion criteria (‘-’ in ***Table 1***) in the CP rule being evaluated. This was different from the final CP algorithm where we dropped the exclusion criteria as discussed below in ***Table 5***.

**Table 4.**
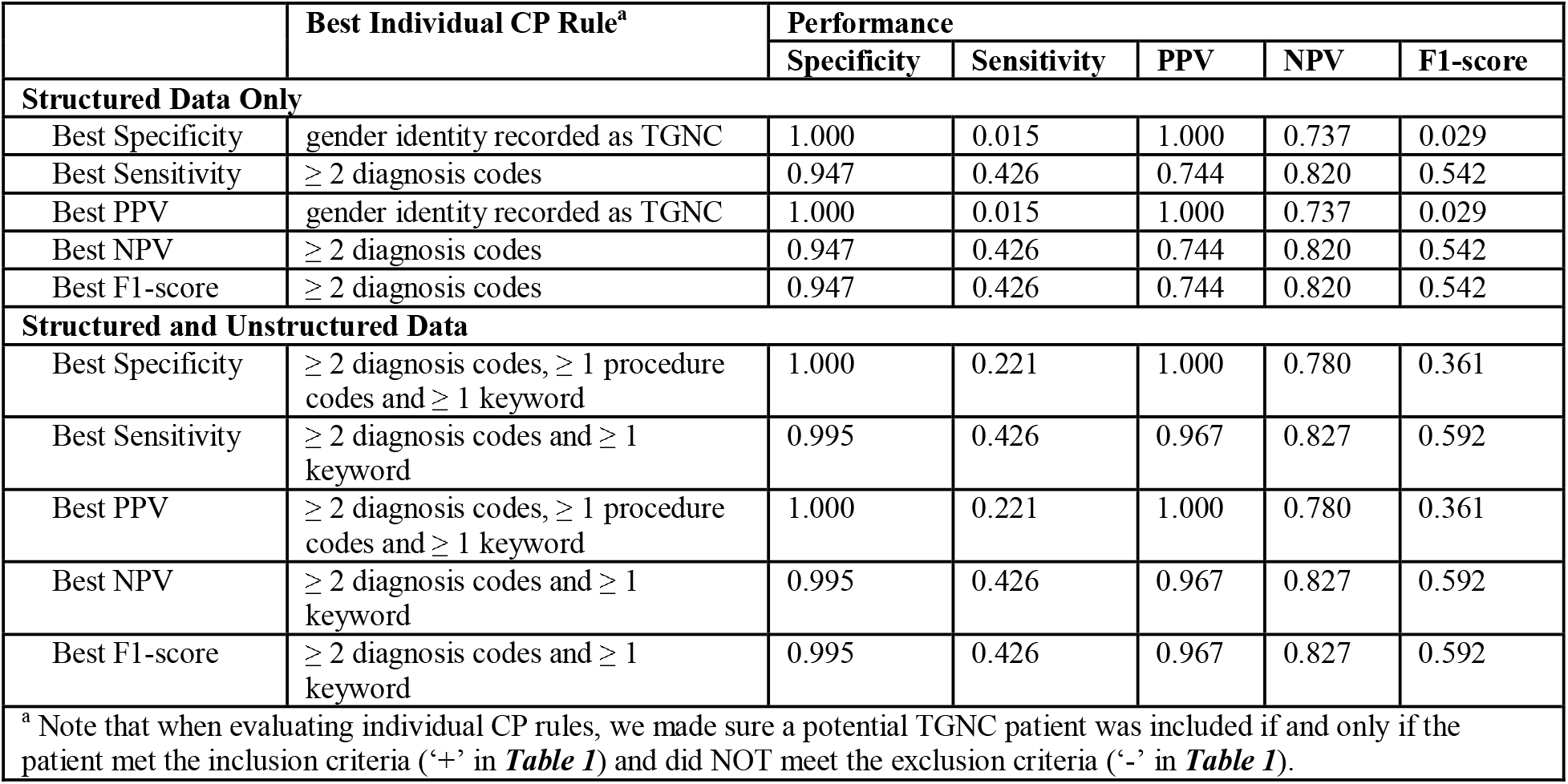
Best performing individual CP rules and their performance.

**Table 5.**
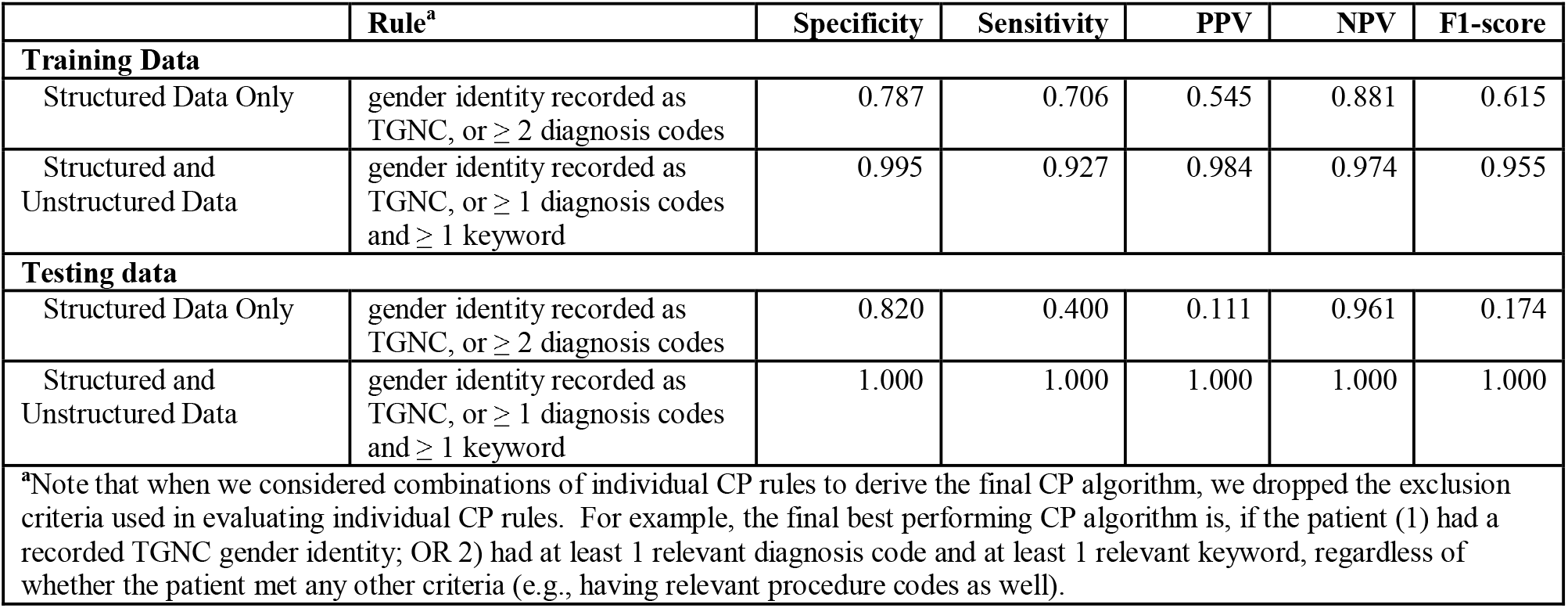
Performance of combined CP rules with the best f1-scores.

Next, we combined individual CP rules to construct the final CP algorithm. To do so, we permutated the different combinations of individual CP rules and evaluated the performance of each combination, considering the same two scenarios (i.e., structured data only vs. both structured and unstructured data). We selected the best performing combinations as the final CP algorithm for identifying TGNC individuals in terms of F1-score. We then measured the performance of the final CP algorithm on the independent testing data. ***Table 5*** shows the experiment results considering the two different scenarios on training and testing data, respectively.

As shown in ***Table 5***, the best performing combination of rules considering structured data only was if the patient: (1) had a recorded TGNC gender identity (i.e., “t*ransgender female / male-to-female*”, “*transgender male / female-to-male*”, “*nonbinary*”, or “*other*”) OR (2) had at least 2 relevant diagnosis records, which had an F1-score of 0.615. When considering both structured and unstructured data, the best performing combination of rules for identifying TGNC patients was if the patient (1) had a recorded TGNC gender identity OR (2) had at least 1 relevant diagnosis code and at least 1 relevant keyword, which had an F1-score of 0.954, a significant improvement over the F1-score in the scenario considering structured data only. When applying the combinations of rules on the testing data: (1) the best performing combination considering structured data only achieved an F1-score of 0.174, while (2) the combination considering both structured and unstructured data achieved a perfect F1-score of 1.0. Using the latter as the final CP algorithm, 1,025 out of the 19,600 potential TGNC individuals were determined to be TGNC.

### Identification of the natal sex of TGNC individuals

Based on Quinn et al, we used the criteria in ***Table 3*** to identify the natal sex of the TGNC individuals as follows:

1. If the patient has a gender identity recorded as “t*ransgender female / male-to-female*”, or “*transgender male / female-to-male*”, the natal sex is determined based on the gender identity record.
2. If the patient has a high precision sex assignment keyword (e.g. “*natal female*”, “*genetic male*”) in notes, the natal sex is determined based on the keyword.
3. If the patient has both MTF and FTM keywords in notes, or relevant procedure/medication records, choose the majority one as natal sex. For example, if the patient has not only a MTF keyword and a MTF procedure, but also has a FTM medication, MTF is assigned as the natal sex.
4. If the patient has both FTM and MTF keywords in notes or both FTM and MTF procedure/medication records, and there is not a clear majority, we choose the natal sex according to the following priority order: procedures, hormonal therapy medications, reassignment keywords, and natal sex keywords.
5. If insufficient information is presented, the patient’s natal sex is assigned as unknown.

Using the 63 TGNC individuals from training set and the 5 TGNC individuals from the testing set, we generated a gold standard dataset through manual chart review. Among the 68 TGNC patients, 27 were MTF and 41 were FTM. Our algorithm identified 29 MTF (5 false positive) and 36 FTM (1 false positive), while 3 TGNC individuals’ natal sex was unknown. **Table 6** shows the performance of the final CP algorithm we developed for determining MTF or FTM status. Among the 1,025 TGNC, 468 was FTM, 403 was MTF, and the remaining 154 had unknown natal sex.

**Table 6.** Performance of the final CP algorithm for determining MTF and FTM status.

|  | Specificity | Sensitivity | PPV | NPV | F1-score |
| --- | --- | --- | --- | --- | --- |
| Transmasculine (FTM) | 0.875 | 0.960 | 0.828 | 0.972 | 0.889 |
| Transfeminine (MTF) | 0.960 | 0.875 | 0.972 | 0.828 | 0.921 |

### TGNC cohort characteristics

We applied the best performing CP algorithms to determine the (1) TGNC status, and (2) MTF/FTM status of the entire potential TGNC cohort extracted from UF Health. **Table 7** shows the demographics of the TGNC cohort compared to the general population in UF Health.

**Table 7.** Demographics of the TGNC patients and general patients in UF Health.

|  | TGNC Patients in UF Health |  |  | General Population in UF Health |  |  |
| --- | --- | --- | --- | --- | --- | --- |
|  | Overall | MTF | FTM | Overall | Male | Female |
|  | N = 1,025<br>(100%) | N = 403<br>(100%) | N = 468<br>(100%) | N = 1,581,490<br>(100%) | N = 727,524<br>(100%) | N = 852,871<br>(100%) |
| **Age** |  |  |  |  |  |  |
| < 3 | 0 (0.0%) | 0 (0.0%) | 0 (0.0%) | 39,849 (2.5%) | 21,160 (2.9%) | 18,669 (2.2%) |
| 3-17 | 264 (25.8%) | 77 (19.1%) | 154 (32.9%) | 266,890 (16.9%) | 140,467 (19.3%) | 126,372 (14.8%) |
| 18-24 | 259 (25.3%) | 98 (24.3%) | 136 (29.1%) | 162,145 (10.3%) | 71,715 (9.9%) | 90,421 (10.6%) |
| 25-64 | 380 (37.1%) | 205 (50.9%) | 160 (34.2%) | 777,845 (49.2%) | 335,112 (46.1%) | 442,689 (51.9%) |
| ≥ 65 | 17 (1.7%) | 15 (3.7%) | 2 (0.4%) | 334,626 (21.2%) | 159,032 (21.9%) | 174,654 (20.5%) |
| Unknown | 105 (10.2%) | 8 (2.0%) | 16 (3.4%) | 135 (0.0%) | 38 (0.0%) | 66 (0.0%) |
| **Race/Ethnicity** |  |  |  |  |  |  |
| Hispanic | 71 (6.9%) | 24 (6.0%) | 36 (7.7%) | 108,540 (6.9%) | 48,090 (6.6%) | 60,446 (7.1%) |
| NHW | 732 (71.4%) | 272 (67.5%) | 354 (75.6%) | 890,187 (56.3%) | 412,577 (56.7%) | 477,594 (56.0%) |
| NHB | 140 (13.7%) | 78 (19.4%) | 43 (9.2%) | 344,048 (21.8%) | 156,267 (21.5%) | 187,774 (22.0%) |
| Other | 82 (8.0%) | 29 (7.2%) | 35 (7.5%) | 238,715 (15.1%) | 110,590 (15.2%) | 127,057 (14.9%) |

NHW = Non-Hispanic White; NHB = Non-Hispanic Black. Age: for TGNC cohort, used age at index; for UFH cohort, used age at October 31, 2019. Note that we included patients aged < 3 in this table for the general population in UF Health.

We descriptively reported the prevalence of 10 chronic conditions in our TGNC cohort in **Table 8** based on the Centers for Medicare & Medicaid Services (CMS)’s Chronic Condition Warehouse (CCW) condition algorithms,^18^ modified for EHR use. We also examined the prevalence of human immunodeficiency virus (HIV) infection in this cohort, as the literature indicated an elevated prevalence of HIV in the TGNC population.

**Table 8.** Prevalence of chronic conditions in the TGNC patients in UF Health.

| Chronic Conditions<sup>a</sup> | General Population in UF Health<sup>b</sup> | TGNC Individuals | Transfeminine (MTF) | Transmasculine (FTM) |
| --- | --- | --- | --- | --- |
|  | N = 1,541,506 (100%) | N = 1,025 (100%) | N = 403 (100%) | N = 468 (100%) |
| ADRD | 28,556 (1.9%) | 11 (1.1%) | 6 (1.5%) | 5 (1.1%) |
| Asthma | 122,652 (8.0%) | 147 (14.3%) | 64 (15.9%) | 58 (12.4%) |
| COPDB | 100,534 (6.5%) | 69 (6.7%) | 41 (10.2%) | 18 (3.8%) |
| **Depression** | **164,768 (10.7%)** | **532 (51.9%)** | **198 (49.1%)** | **253 (54.1%)** |
| Diabetes | 144,563 (9.4%) | 90 (8.8%) | 51 (12.7%) | 26 (5.6%) |
| Hypertension | 342,098 (22.2%) | 213 (20.8%) | 109 (27.0%) | 75 (16.0%) |
| Breast cancer | 19,601 (1.3%) | 11 (1.1%) | 4 (1.0%) | 5 (1.1%) |
| Colorectal cancer | 10,916 (0.7%) | 4 (0.4%) | 3 (0.7%) | 1 (0.2%) |
| Prostate cancer | 18,131 (1.2%) | 2 (0.2%) | 1 (0.2%) | 0 (0.0%) |
| Lung cancer | 11,776 (0.8%) | 4 (0.4%) | 3 (0.7%) | 1 (0.2%) |
| **HIV** | **8,062 (0.5%)** | **70 (6.8%)** | **57 (14.1%)** | **4 (0.9%)** |

<sup>a</sup>ADRD: Alzheimer's Disease and Related Disorders or Senile Dementia; COPDB: Chronic Obstructive Pulmonary Disease and Bronchiectasis; HIV: Human immunodeficiency virus infection <sup>b</sup>We excluded patients age < 3 or Unknown from the general population in UF Health in this table.

### Prevalence of TGNC and TGNC subgroups

We calculated the prevalence of confirmed TGNC individuals and subsequently TGNC with MTF/FTM status from 2012 to 2019 (up to October 2019). For each eligible TGNC, the date of the first visit associated with the TGNC status was considered the index date. To be included in the numerator for a given calendar year, a patient had to have at least one encounter in the UF Health system at any time during that year and have an index date that was in or before that year. The denominator comprised of all patients who had at least one encounter in the UF Health system during the same year. Each prevalence rate was accompanied by a 95% confidence interval (CI) calculated based on the Fleiss quadratic correction using the OpenEpi statistical calculator.^19^ All prevalence rates and the corresponding 95% CIs were expressed as per 100,000 persons, as shown in **Table 9** (see next page).

**Table 9.** The unadjusted prevalence of TGNC and TGNC subgroups in UF Health from 2012 to 2019.

|  | TGNC | Total Patients | Prevalence estimate (95% CI)<sup>a</sup> | MTF | Total Natal Males | Prevalence estimate (95% CI)<sup>a</sup> | FTM | Total Natal Females | Prevalence estimate (95% CI)<sup>a</sup> |
| --- | --- | --- | --- | --- | --- | --- | --- | --- | --- |
| 2012 | 24 | 374,262 | 6.4<br>(4.3 - 9.5) | 14 | 158,059 | 8.9<br>(5.3 - 14.9) | 9 | 216,203 | 4.2<br>(2.2 - 7.9) |
| 2013 | 62 | 396,466 | 15.6<br>(12.2 - 20.0) | 40 | 168,141 | 23.8<br>(17.5 - 32.4) | 16 | 228,290 | 7.0<br>(4.3 - 11.4) |
| 2014 | 85 | 433,011 | 19.6<br>(15.9 - 24.3) | 56 | 184,443 | 30.4<br>(23.4 - 39.4) | 25 | 248,495 | 10.1<br>(6.8 - 14.9) |
| 2015 | 149 | 485,741 | 30.7<br>(26.1 - 36.0) | 79 | 208,750 | 37.8<br>(30.4 - 47.2) | 55 | 276,941 | 19.9<br>(15.3 - 25.9) |
| 2016 | 249 | 528,945 | 47.1<br>(41.6 - 53.3) | 125 | 228,486 | 54.7<br>(45.9 - 65.2) | 108 | 300,414 | 36.0<br>(29.8 - 43.4) |
| 2017 | 421 | 512,887 | 82.1<br>(74.6 - 90.3) | 181 | 219,589 | 82.4<br>(71.3 - 95.3) | 212 | 293,235 | 72.3<br>(63.2 - 82.7) |
| 2018 | 547 | 530,732 | 103.1<br>(94.8 - 112.1) | 225 | 226,347 | 99.4<br>(87.2 - 113.3) | 284 | 303,958 | 93.4<br>(83.2 - 104.9) |
| 2019<sup>b</sup> | 616 | 500,419 | 123.1<br>(113.8 - 133.2) | 248 | 213,095 | 116.4<br>(102.8 - 131.8) | 321 | 286,960 | 111.9<br>(100.3-124.8) |
| 2012-2019 | 1025 | 1,541,506 | 66.5<br>(62.6 - 70.7) | 403 | 706,326 | 57.1<br>(51.8 - 62.9) | 468 | 834,136 | 56.1<br>(51.3 - 61.4) |

<sup>a</sup>Per 100,000 population. <sup>b</sup>Only available up to October 31, 2019.

## Discussion and conclusion

In this study, we developed and validated a computable phenotype algorithm for identifying TGNC individuals in EHRs using both structured (discrete fields, and diagnosis and procedure codes) and unstructured (clinical notes) data. Our results showed that the best performing CP algorithm achieved a perfect F1-score on the independent testing data.

Our study extended the work conducted by Quinn et al in several important ways. First, we considered the discrete gender identity field that is being increasingly adopted in modern EHR systems and expanded the diagnosis codes to include ICD-10-CM codes in addition to ICD-9-CM codes. Second, we significantly expanded the list of keywords related to TGNC status. The keyword expansion improved the sensitivity of our CP algorithm and led to the identification of significantly more TGNC individuals. Third, and perhaps more importantly, our CP algorithm is automated and does not require manual chart review of uncertain cases, where Quinn et al manually reviewed thousands of uncertain cases to determine their TGNC status. Even though our CP algorithm is not perfect, the high performance (i.e., 0.955 and 1.0 F1-score on the training and testing data, respectively) minimizes the bias introduced by misclassification errors for downstream analyses. Lastly, as Quinn et al used Kaiser Permanente internal codes, their approach is not generalizable to other health systems. In contrast, our final CP algorithm is simple (i.e., “*gender identity recorded as TGNC, or* ≥ *1 diagnosis codes and* ≥ *1 keyword*”), does not rely on any internal knowledge of the health system, and thus generalizable to other health systems (e.g., other partners in OneFlorida and PCORnet).

Overall, the unadjusted prevalence of TGNC individuals was estimated to be 97.7 (95% CI: 89.9 - 106.2) per 100,000 or 0.10% in 2018, and 117.2 (95% CI: 108.3 - 126.8) per 100,000 or 0.12% in 2019, in the UF Health EHR system. There was also a clear increasing trend in the proportion of population identified as TGNC in the past few years. Our prevalence estimates were slightly lower than the previously reported rate of 0.66% TGNC adults in Florida based on telephone surveys conducted as part of the Centers for Disease Control and Prevention’s Behavioral Risk Factor Surveillance System (BRFSS).^20^ In a meta-analysis of population surveys, a slightly lower prevalence of TGNC individuals (390 per 100,000 or 0.39%) was reported among US adults.^21^ Noting that significant barriers, such as stigma and discrimination, may impact TGNC individuals’ desire and ability to access appropriate care,^22,23^ the prevalence of TGNC individuals in EHR data is expected to be lower than that in survey data. On the other hand, our TGNC prevalence estimates were significantly higher than those reported in Quinn et al. By 2014, the prevalence rates of TGNC individuals reported in Quinn et al were 38 (95% CI: 32 - 45), 44 (95% CI: 42 - 46), and 75 (95% CI: 72 - 78) per 100,000 in the three separate Kaiser Permanente health plans. One potential reason for the higher rates observed in the UF Health EHR system is that our approach identified and included significantly more keywords related to TGNC status, and a large proportion of TGNC individuals were identified in clinical notes using keywords.

The distribution of prevalence rates by age group observed in our TGNC cohort was consistent with that reported in national surveys. In our 2019 data, the prevalence rate of TGNC individuals was 0.15%, 0.39%, 0.096%, and 0.015% in the 3-17 year-olds, 18-24 year-olds, 25-64 year-olds, and adults older than 65, respectively. In the BRFSS data, the prevalence rate of TGNC individuals was 0.75%, 0.67%, and 0.55% in the 18-24 year-olds, 25-64 year-olds, and adults older than 65, respectively. In both data sources, the prevalence of TGNC individuals decreased as age increased, and the highest prevalence was observed in the 18-24 year-olds. Both population surveys and RWD from EHR systems are important data sources for understanding the unique health burdens in the TGNC population. These data sources complement each other as they may capture different TGNC subpopulations. More accurate estimates of the TGNC population and subpopulation sizes could be derived if both data sources are considered.

The prevalence rates of the chronic conditions and HIV infection observed in our TGNC cohort were consistent with those reported in the literature. Compared to the overall UF Health population, the TGNC individuals had significantly higher rates of depression (51.9% vs. 10.7%) and HIV (6.8% vs. 0.5%).^24,25^ Further, the prevalence of HIV was significantly higher among the transfeminines than the transmasculines (14.1% vs. 0.9%). This disparity in HIV prevalence was often explained by the difference in risky behaviors between the two groups. Nevertheless, evidence gaps remain for contextual factors specific to the transgender experience.^25^

Our study is not without limitations. First, our CP algorithm only considers the existence of certain TGNC relevant keywords but does not take into consideration the contexts in which the keywords are used. For example, our CP algorithm would not be able to account for negations (e.g., “*he does not consider himself a transgender*”) or references to people other than the patients themselves (e.g., “*he lived with a transgender relative*”). Our CP algorithm would also not be able to account for misuse of TGNC terms by physicians. For example, in one note, the patient was stated as “*a male who is trans female (born female living as male) and currently taking testosterone cypionate for male hormone*”, where the correct technical term should be “*trans male*” (i.e., a man who was assigned female at birth) rather than “*trans female*.” From a CP algorithm perspective, the contexts in which the keywords are used can be explored using more advanced natural language processing (NLP) methods (e.g., negation detection). Often times, using NLP methods requires significant amount of effort but results in a small or moderate improvement in CP performance. Nevertheless, whether to consider advanced NLP methods when developing a CP algorithm will be based on the specific downstream application needs. Second, as shown in our study, CP algorithms are not static and regular refinements of the CP algorithms are needed to keep them up to date (e.g., transition from ICD-9 to ICD-10). As a best practice for developing and using CP algorithms, local validation and refinement should always be performed.

## Data Availability

Due to the nature of this research, participants of this study did not agree for their data to be shared publicly, so supporting data is not available.

## Acknowledgement

This work was supported in part by NIH grants UL1TR001427, 1R01CA246418, R21CA245858, U18DP006512, and PCORI grant ME-2018C3-14754. The content is solely the responsibility of the authors and does not necessarily represent the official views of the NIH and PCORI.

